# Combining Navigator and Optical Prospective Motion Correction for High-Quality 500 μm Resolution Quantitative Multi-Parameter Mapping at 7T

**DOI:** 10.1101/2021.10.26.21265506

**Authors:** Lenka Vaculčiaková, Kornelius Podranski, Luke J. Edwards, Dilek Ocal, Thomas Veale, Nick C. Fox, Rainer Haak, Philipp Ehses, Martina F. Callaghan, Kerrin J. Pine, Nikolaus Weiskopf

## Abstract

**PURPOSE:** High-resolution quantitative multi-parameter mapping shows promise for non-invasively characterizing human brain microstructure but is limited by physiological artifacts. We implemented corrections for rigid head movement and respiration-related B0-fluctuations and evaluated them in healthy volunteers and dementia patients.

**METHODS:** Camera-based optical prospective motion correction (PMC) and free-induction decay (FID) navigator correction were implemented in a gradient and RF-spoiled multi-echo 3D gradient echo sequence for mapping proton density (PD), longitudinal relaxation rate (R1) and effective transverse relaxation rate (R2*). We studied their effectiveness separately and in concert in young volunteers and then evaluated the navigator correction (NAVcor) with PMC in a group of elderly volunteers and dementia patients. We used spatial homogeneity within white matter (WM) and gray matter (GM) and scan-rescan measures as quality metrics.

**RESULTS:** NAVcor and PMC reduced artifacts and improved the homogeneity and reproducibility of parameter maps. In elderly participants, NAVcor improved scan-rescan reproducibility of parameter maps (coefficient of variation decreased by 14.7% and 11.9% within WM and GM respectively). Spurious inhomogeneities within WM were reduced more in the elderly than in the young cohort (by 9% vs 2%). PMC increased regional GM/WM contrast and was especially important in the elderly cohort, which moved twice as much as the young cohort. We did not find a significant interaction between the two corrections.

**CONCLUSION:** Navigator correction and PMC significantly improved the quality of PD, R1 and R2* maps, particularly in less compliant elderly volunteers and dementia patients.

## INTRODUCTION

Quantitative magnetic resonance imaging (qMRI) offers the possibility to infer upon human brain microstructure non-invasively (1,2), using links between MR parameters and brain tissue components. The time-efficient multi-parameter mapping (MPM) protocol (3,4) accurately and robustly maps the qMRI parameters: longitudinal relaxation rate (R1), effective transverse relaxation rate (R2*), proton density (PD) and magnetization transfer (MT) saturation - parameters which are sensitive and (relatively) specific to myelin and iron tissue components in the brain (5,6).

Characterization of small anatomical structures requires, apart from high tissue specificity, high spatial resolution and low level of artifacts. Moving to ultra-high field such as 7T allows for reduction of voxel size as the signal to noise ratio (SNR) increases with field strength (7). But at this higher image resolution, correction of head motion becomes crucial. Additionally, 7T MRI brings challenges such as flip angle variations across the brain due to poorer B1 homogeneity (8) and increased physiological artifacts (9,10). While B1 inhomogeneity artifacts can to a large extent be corrected for using B1 mapping (11), motion and physiological artifacts still limit high-resolution ultra-high field MRI. Here we propose and test a combination of methods to overcome these artifacts.

A broad range of studies can benefit from correction of physiological artifacts and the consequent improved reliability of parameter mapping. It is especially important in studies addressing questions related to thin layers in the neocortex (12,13) or small subcortical nuclei (14). Similarly, studies on gray and white matter (GM, WM) maturation (15-17), aging (18,19) or trauma-related changes (20) indicate small variations in relaxation parameters and myelin sensitive metrics on the order of a few percent only. Therefore, physiological artifacts need to be minimized to be able to detect such small effects of interest. This becomes even more important in certain groups of patients or volunteers; for example physiological dynamic field fluctuations are higher in cohorts of dementia patients (21,22).

Head motion during data acquisition reduces the effective resolution (23) and manifests as image artifacts (24,25). Prolonged acquisition times (TA), often a necessity in high resolution imaging, can lead to participant discomfort and increased involuntary movements. Cardiac pulsations, swallowing or breathing also contribute to the deterioration of image quality (26). Moreover, motion during data acquisition increases with age (27), while patients with cognitive impairment may even have difficulty complying with or remembering instructions to lie still. Motion artifacts such as blurring and ghosting can be reduced, for example, via shortening TA using parallel imaging (28,29), opting for acquisition strategies that are inherently more robust to motion (30), using self-navigated methods (31), navigator echoes (32), fat navigators (33,34) or external tracking devices (35). Information on the head position can be used for correction either prospectively or in the post-processing after data acquisition. In this study, we applied an optical prospective motion correction (PMC) system to track and correct for rigid head motion. The optical system (36) had been previously evaluated in relation to qMRI at 3T (37) as well as qualitative MRI (23) and quantitative susceptibility mapping at 7T (38).

Subject motion within as well as outside of the imaged region along with physiology-related processes gives rise to dynamic B0 field fluctuations (39-42). The major contributor to time-varying fields is respiration, when lungs periodically fill with air that has different magnetic susceptibility than the surrounding tissue (43). The moving chest changes the distribution of the B0 field, therefore violating the assumption of static magnetic field homogeneity for accurate spatial encoding, even as far away as in the brain (9). There are several ways to correct breathing-induced frequency shifts, such as modulation of the B0 shims in real time either by monitoring chest position (10), the B0 field using NMR field probes (35,44,45), or tracking the perturbing fields with phase navigators and incorporating these in the image reconstruction (21,46). We opted for free induction decay (FID) navigator correction (NAVcor), since it does not require extra hardware and is time efficient as a result of not encoding the navigators in space.

For this study, we implemented FID navigators and PMC corrections into a custom 3D gradient-recalled echo sequence (GRE) (47). We developed a novel FID navigator estimation method, which is robust to low SNR and outliers in the FID signal. We investigated the effectiveness of PMC and temporal B0-fluctuation corrections separately and their combination for improving the quality of high-resolution quantitative parameter maps in young healthy volunteers. In another experiment we tested the combination of both PMC and navigator correction in a group of elderly participants, some of whom suffered from progressive neurodegenerative diseases, specifically Alzheimer’s disease (AD) and posterior cortical atrophy (PCA) (48) which is an atypical (“visual”) variant of AD where the posterior cortex is focally and disproportionately affected. Both qualitative visual assessment and quantitative quality metrics of spurious spatial inhomogeneities and scan-rescan reproducibility were used to determine the effectiveness of the correction methods.

## METHODS

### Optical PMC system

The PMC system (KinetiCor, HI, USA) (36,37) used an in-bore optical camera for tracking the position of a Moiré phase marker at a frame rate of 80 Hz. The camera inferred the position of a phase marker which is coupled with the head position, based on the combination of fixed markings and an optical Moiré pattern. It measures six degrees of freedom - three translational and three rotational, all at a very high precision (36). A calibrated affine transform between camera and scanner coordinate system was used to dynamically translate the marker position into the imaging field of view (FOV) position (49) and the information was logged in relation to the initial pose at the start of each scan. The FOV was adjusted every repetition time (TR) based on the latest position without any temporal filtering of the head position time course.

### Imaging pulse sequence

The PMC functionality and navigator-based magnetic field monitoring were implemented in a custom-made gradient and RF-spoiled multi-echo 3D GRE sequence with alternating readout gradient polarity using the vendor’s programming framework (Siemens IDEA VB17; Figure 1). The PMC system updated imaging gradients and frequency and phase of the RF pulse once every TR. The position was updated 2 ms before the end of each TR and the RF excitation of the next TR (Figure 1).

**FIGURE 1.**
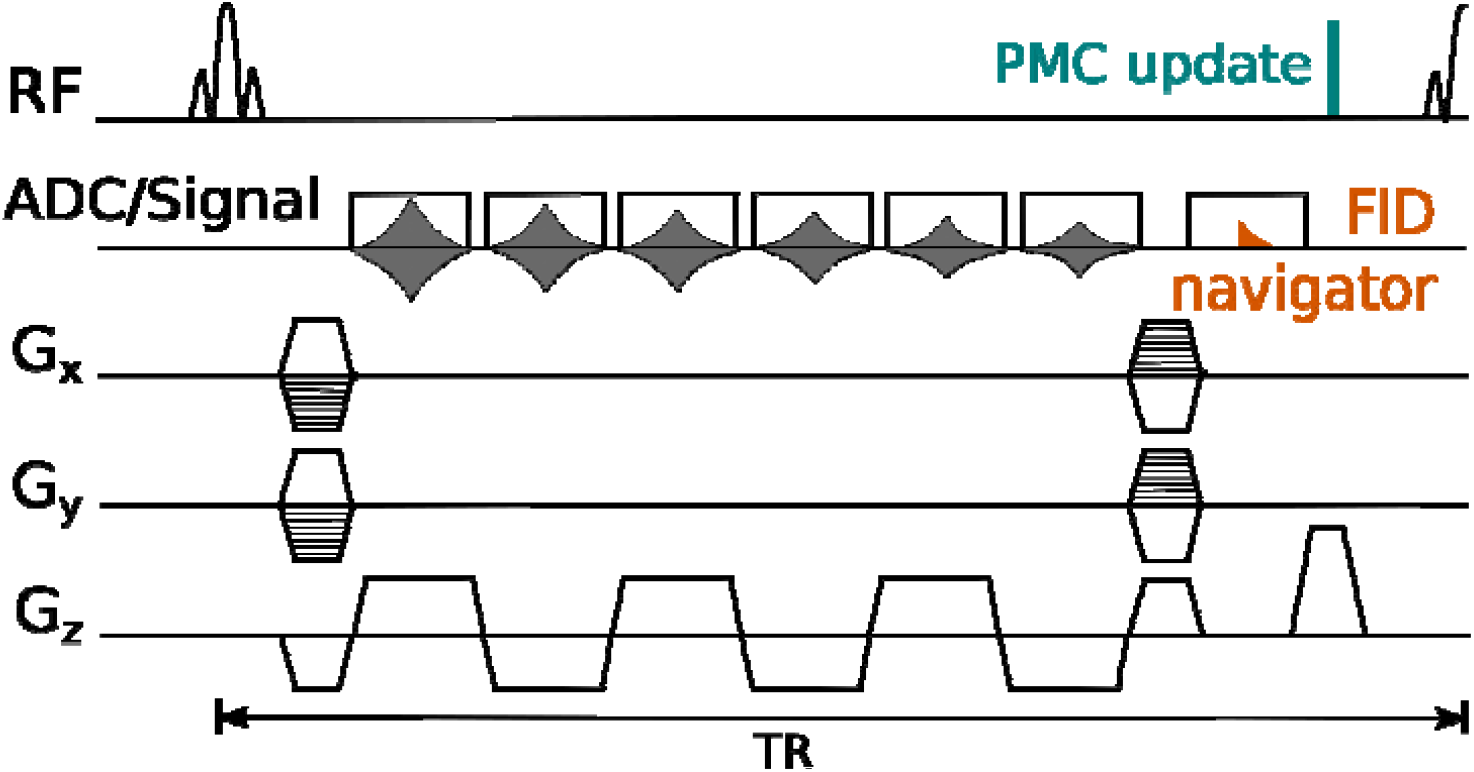
Pulse sequence diagram of a conventional 3D multi-echo GRE sequence using a non-selective RF sinc pulse for excitation and bipolar readouts, modified to include B0-fluctuation monitoring (orange) and motion correction (turquoise). Only part of each acquired FID navigator signal was used. The selection is a trade-off between SNR (i.e. using as many early FID points as possible) and reducing sensitivity to nuisance eddy currents from the readout and rewinder gradients (selected signal highlighted in orange).

The modifications of a standard 3D multi-echo gradient sequence required to monitor temporal B0-field fluctuations involved a navigator data acquisition in the center of k-space. In order to increase sensitivity to phase evolution and not to extend the minimum readout echo time (TE), navigator sampling was placed after the imaging echoes. The temporal resolution of the field evolution monitoring was equal to TR. The first ca. 500 μs of the navigator following the end of the rephasing gradient lobe were discarded due to residual eddy currents and the subsequent 72 μs were used to estimate frequency shift (orange signal in Figure 1).

### Participants and experimental setup

Ten younger healthy (mean age 30, standard deviation [SD] 4 years) and ten elderly volunteers and patients (4*x* AD, 3*x* PCA and 3*x* control group with mean age 61, SD 4 years) were scanned on two 7T Magnetom whole body MR systems (Siemens Healthineers, Erlangen, Germany) at the MPI-CBS and the Wellcome Centre for Integrative Neuroimaging, University of Oxford, respectively. Both were equipped with a circularly polarized single-channel transmit and 32-channel RF receive head coil (Nova Medical, Wilmington, MA, USA). Written informed consent was obtained before scanning. The studies on the younger volunteers and elderly/patient groups were approved by the Ethics Committee of the Medical Faculty of the University of Leipzig and the London-Harrow Research Ethics Committee, respectively. Dielectric pads were placed over the temporal lobes to counter B1 field inhomogeneity (50). Head movement was detected using the PMC system, with the Moiré marker attached onto a mini mouthguard molded to the participant’s upper teeth (similar to (51)). The marker was affixed to the mouthguard via a plastic extension whose length was adjusted to center the marker within the FOV of the tracking camera outside the transmit coil. Participants were instructed to stay still during data acquisition and not to bite on the mouthguard. In the younger volunteer group only, pulse and respiration were monitored and recorded using an MP150 system (BIOPAC Systems, Goleta, CA). Throughout the experiment, the younger cohort watched a movie (without sound), projected onto a screen that was viewed via a mirror mounted onto the RF coil.

### Experimental designs

In the first study in the younger cohort, data were measured with the PMC turned on and off (*PMC on* and *PMC off* conditions) in a single session, but navigators were acquired in each measurement. Each dataset was reconstructed with and without retrospective B0-fluctuation correction (*NAVcor on* and *NAVcor off* conditions). This resulted in a 2 × 2-factorial design, which allowed the assessment of the PMC and NAVcor corrections separately, together and their interaction. The second study in elderly healthy volunteers and dementia patients was designed for an assessment of scan-rescan reproducibility for *NAVcor on* versus *NAVcor off*. The data were acquired in two separate sessions, always under the *PMC on* condition and reconstructed for both NAVcor conditions.

### Data acquisition

At the beginning of each session, a low resolution RF transmit/B1^+^ map was measured to adjust the transmitter voltage for optimal imaging of the occipital lobe in each individual, since the developments targeted studies of the visual system. Then, vendor’s automatic shimming was iterated 3 times and calibration measurements of RF transmit and static magnetic fields were acquired (3,11). Two whole brain GRE scans at 500 µm isotropic resolution followed, with predominant PD and T1 weightings, as per the MPM protocol (3). Non-selective RF sinc pulses with bandwidth-time product of 6, flip angles and durations of α_PDw_ = 8°, 182 μs (PD-weighting) and α_T1w_= 30°, 680 μs (T1-weighting) were used for excitation. FOV dimensions were 248×217×176 mm^3^, 496 × 434 × 352 matrix size (read [head-foot] x phase [anterior-posterior] x [left-right; fast/inner phase encoding direction]) and its orientation was sagittal with 30° in-plane rotation (pitched anteriorly upwards). 6 echoes were equispaced between 2.8 ms and 16.1 ms with readout bandwidth of 420 Hz/pixel. TR was 27.5 ms, which was 2.7 ms longer than what could be achieved without using navigators. Factor 2×2 parallel imaging was used in the phase encoding directions. TA for a single GRE scan was 18 min 13 sec.

The protocols between the first and the second study slightly differed due to differing acquisition time restrictions and further reduction of bias due to undesired magnetization transfer effects (52). The following parameters differed in the second study: α_T1w_= 25°; RF durations - 140 μs (PD-weighting),1368 μs (T1-weighting); equispaced TEs between 3.22 ms and 16.52 ms; TR = 28.5 ms (2.8 ms longer than the minimum TR without navigators); TA = 20 min 52 sec.

### B0-correction and image reconstruction

While motion correction was applied prospectively, temporal B0-fluctuations were estimated from the navigator acquisitions and corrected retrospectively as follows. For each TR one navigator dataset was acquired consisting of 992 data points with a dwell time of 2.4 μs. Only a subset of 30 data points (i.e. 72 μs long) starting 492 µs from the last gradient activity was selected for further processing, to reduce eddy current effects and minimize low SNR issues. For each TR the B0 and corresponding frequency (f0) change were estimated in a series of steps detailed below and in the navigator processing section of Figure 2.

**FIGURE 2.**
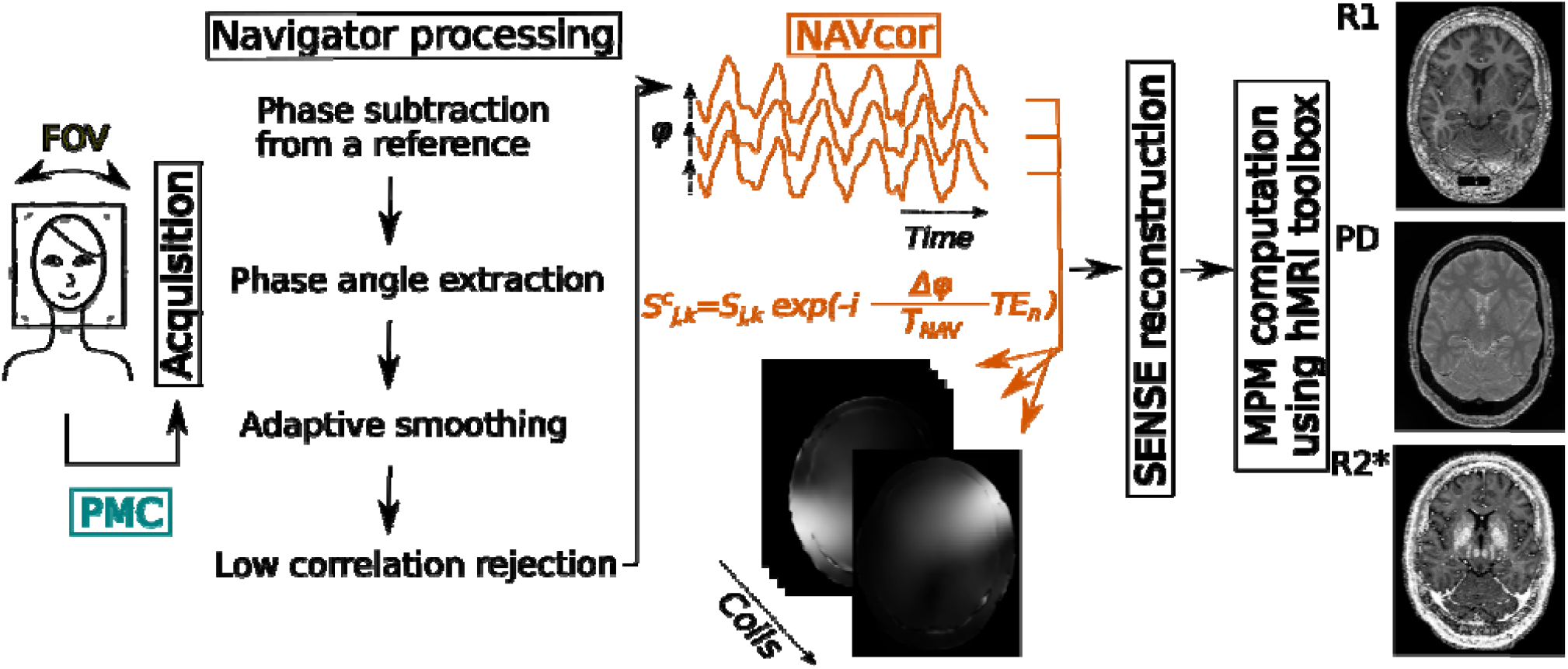
Schematic of the placement of PMC (turquoise) and NAVcor (orange) within the image acquisition, reconstruction and data processing pipeline (data flow from left to right). While motion correction was prospective and applied during data acquisition (left column, turquoise), navigator correction was applied retrospectively. The navigator data was processed to minimize outliers and artifacts using an adaptive smoothing and consistency analysis. The resulting dynamic frequency Δf0 and phase estimates Δφ were used to correct acquired signal S at phase encoding positions j, k, where T_NAV_ is the time between RF pulse and FID navigator acquisition. Both imaging and auto-calibration reference k-space lines were corrected for phase errors introduced by ΔB0 per channel and for each TE_n_ where n=1…6, with the assumption that Δf0 did not change over the time the 6 echoes were acquired. Following navigator correction, images were reconstructed using an in-house implementation of 2D SENSE. Quantitative maps were computed from the different weighted images and echoes using the hMRI toolbox.

The following steps were performed for each RF receive channel separately. The phase of each complex-valued navigator data point was subtracted pointwise from that of the corresponding data point of the navigator acquired for the tenth TR (selected to be the phase reference). The median phase φ was taken over the 30 FID data points such that φ(n)=f0(n).2π.T_nav_ should represent the temporal fluctuation of f0 at the n-th TR and T_nav_ the time between the RF excitation and the acquisition of the FID navigator (19.0/19.4 ms for the younger/elderly cohort).

Due to the rather long T_nav_, some points in the time-series of φ sporadically suffered from low SNR and outliers. While outliers can cause individual lines to have incorrect phase, they can also cause phase unwrapping to fail propagating error to all subsequent lines. Therefore, the time series of φ underwent additional conditioning. In order to balance preserving the temporal features of the navigator time courses with minimizing the impact of outliers due to low SNR, regularization based on SNR was introduced. The fundamental idea was to generate a regularized time series by mixing the original time series with a highly smoothed version free of outliers. The relative mixing weights were adjusted for each time point based on its SNR. Thus, for high SNR the original values would be preserved, but for extremely low SNR they would be effectively replaced by the value from the highly smoothed time course.

The SNR was estimated for each time point of each navigator time course as the mean divided by the standard deviation of the magnitude of the complex-valued 30 data points per TR. Low SNR time periods were characterized as a segment of the navigator trace below a specified SNR threshold for a certain minimum number of consecutive TRs (see below for typical cut-off values). The highly smoothed version of the original navigator trace φ was constructed as an artifact free reference. The original time series underwent a smoothing step based on a convolution with a cosine lobe window.

The three free parameters of the regularized time course estimation (smoothing filter window for the reference time course, SNR threshold, low SNR detection window size) were simultaneously and automatically optimized for each RF channel. The filter window size was allowed to vary from ca. 5 mins to the full time series length of ca. 20 mins, the SNR threshold from 40-60 and the low SNR detection window size from 4-25 TRs (range of plausible parameters determined by visual inspection of problematic raw navigator traces), until large discontinuities (reflecting unwrapping failures) were no longer detected in the time-series. This was confirmed by checking that the difference between the minimum and maximum of the trace was less than 3.5π (∼90 Hz) and also that there was not an increase of more than 1.5π (∼40 Hz) within any 30 s long segment. The set of lowest parameter values, which fulfilled these criteria for all channels, was used for all RF receive channels of the respective measurement, i.e., the lowest effective window size, SNR threshold and SNR detection window size were used for further processing.

Finally, channels with a poor correlation to the rest of the channels (defined as traces whose correlation coefficient after smoothing with a sliding window size of 250 TRs, approximating one respiratory cycle, was below 0.5) were replaced with a modeled trace created as a pointwise mean of the well-correlated channels.

The estimated dynamic frequency changes (Δf0(n)= f0(n)-f0(10)) were then used for correcting every k-space line on a per RF channel and per TE basis as shown in NAVcor section of Figure 2. Briefly, the channel- and echo-specific dynamic phase shift was predicted from the estimated dynamic frequency change Δφ(n) (= Δf0(n).2π.T_nav_), and corrected by multiplying the original signal with the inverse complex phase factor (Figure 2, center).

Navigator correction was applied to all k-space lines - irrespective of whether they were acquired as an auto-calibration reference k-space line for reconstructing sensitivity maps or for reconstruction of the final image. Navigator correction was performed as the first step before the image reconstruction, i.e. prior to coil sensitivity matrix estimation using an in-house MATLAB implementation (MathWorks, MA) of 2D SENSE (28) reconstruction with auto-calibration inspired by mSENSE (53). All datasets were reconstructed both with (*NAVcor on*) and without (*NAVcor off*) navigator correction.

### Parameter maps computation

Reconstructed PD- and T1-weighted images were coregistered for each participant using SPM12 (http://www.fil.ion.ucl.ac.uk/spm/). Then, R1, PD and R2* maps were estimated using custom-made MATLAB tools including the hMRI toolbox (https://hMRI.info) (54) implemented in the SPM12 framework (Figure 2). Based on a combination of all echoes from the PD- and T1-weighted images, R2* maps were estimated using the weighted ordinary least squares fit utilizing both contrasts. PD maps were corrected for R2* bias by extrapolating the signal to TE=0 (55) and calibrated to 69 p.u. of WM values in voxels exceeding tissue probability of 95% (56). Parameter maps were corrected for geometric gradient nonlinearity distortions using the gradunwarp toolbox https://github.com/Washington-University/gradunwarp) (57,58).

### Data quality assessment: navigators, motion, homogeneity and scan-rescan reproducibility

General data quality and plausibility checks included the estimation of the averaged integrated motion metric (59). We assessed the quality of the parameter maps by visual inspection and quantitatively. LV visually assessed all parameter maps with a special focus on artifacts driven by motion and B0 changes, such as ringing, ghosting, blurring and smooth changes in parameter values. Quantitative assessment consisted of estimating the level of spurious spatial inhomogeneities in the parameter maps and the scan-rescan coefficient of variation (CoV).

In the younger cohort, we employed an inhomogeneity metric for the quantitative assessment, since repeated measurements were not available. GM and WM exhibit relatively constant PD, R1 and R2* values across the brain. In particular, the biological spatial variability is typically smaller than the inhomogeneities caused by image artifacts. As an estimate of spatial inhomogeneity, we determined the inhomogeneity CoV, which was defined as the standard deviation of the parameter values across all voxels in a region of interest (ROI) divided by the mean parameter value across all voxels in the same ROI. GM and WM ROI masks were based on tissue probability masks created by segmentation in SPM during quantitative MPM computation and further gradient nonlinearity-corrected to match the parameter maps. The masks were thresholded at tissue probabilities of 0.95 and 0.97 for GM and WM respectively and eroded by 2 voxels to achieve conservative masks and avoid partial voluming with other tissue types. The inhomogeneity CoV was determined for all 2×2 acquisition conditions (PMC on/off and NAVcor on/off) and parameter maps (PD, R1, R2*). A two-way ANOVA was performed to determine main effects of and interaction between PMC and navigator correction separately in PD, R1 and R2* maps (significance threshold of p < 0.05).

For the scan-rescan experiment in the elderly and patient cohorts, we determined, in addition to the homogeneity CoV the scan-rescan reproducibility by determining the scan-rescan CoV computed as the standard deviation for each voxel across the two measurements divided by the mean (4). The mean scan-rescan CoV was calculated for the GM and WM regions as described above. We tested for a significant improvement due to the navigator correction by one-tailed t-test on the scan-rescan CoV data (p < 0.05).

### Code availability statement

Source code for navigator correction and data processing can be downloaded from the project repository on GitHub: https://github.com/lenkavaculciakova/NavCorPMC.

## RESULTS

### Qualitative assessment of navigators

Navigator traces were compared to measurements with a respiration sensor as well as the PMC system in the younger cohort, in order to identify common factors. Figure 3 shows a typical respiration trace, navigator time course and translational and rotational motion traces. All techniques can detect the dynamic breathing patterns via different mechanisms: mechanic activation of respiration sensors attached to the chest; susceptibility-induced dynamic field changes and rigid body motion of the head due to respiration. Motion accompanied with a changed respiration pattern is observed both in the motion traces and navigators.

**FIGURE 3.**
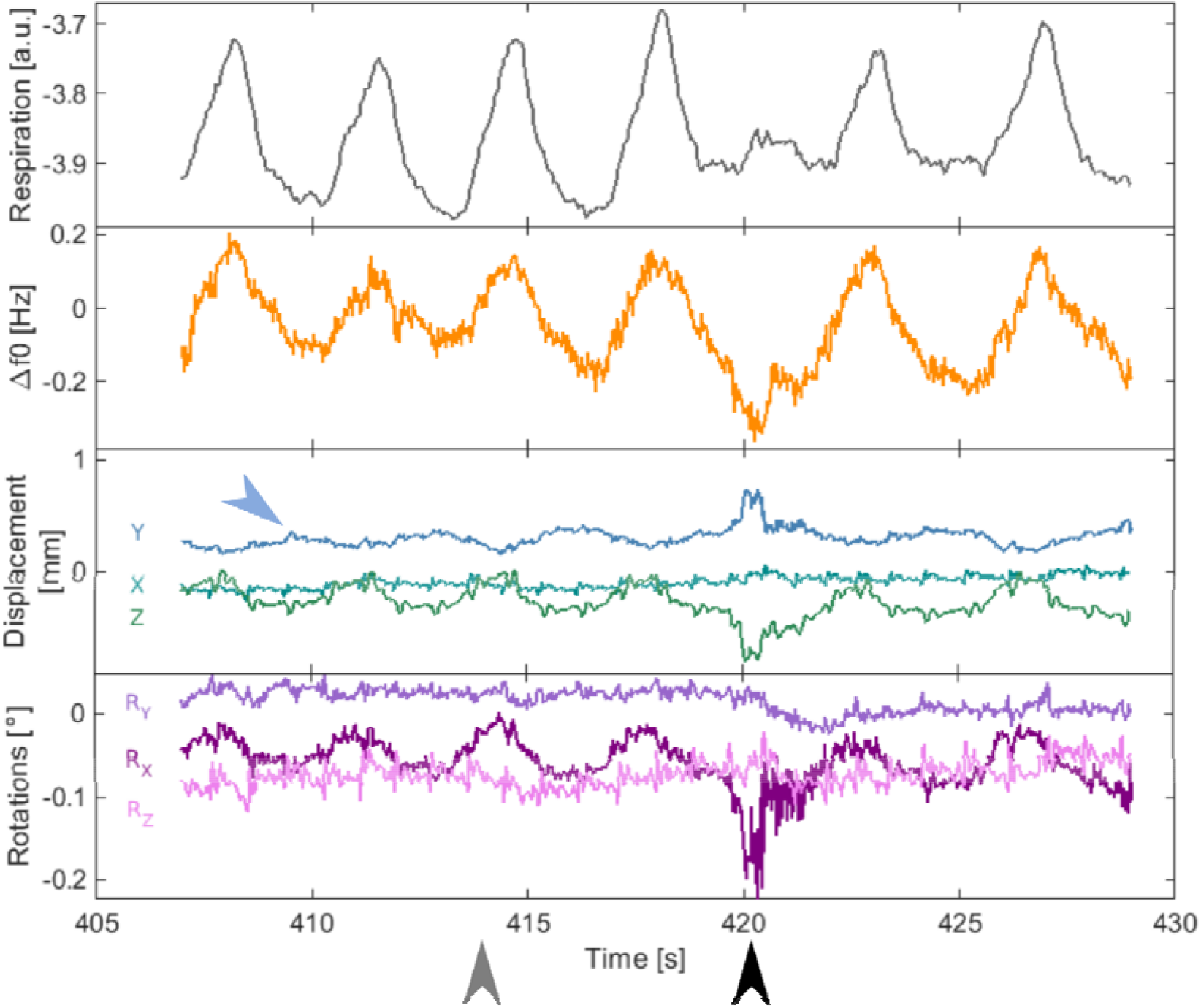
Synchronized time courses from a respiration sensor (gray, top row), navigator (orange, second row) and the PMC system - translational (green/blue, third row) and rotational head motion (purple, bottom row). Each trace type detected subject respiration (e.g., gray arrow points to breathing in motion) and also other types of motion (black arrow). Additionally, cardiac pulse related motion was picked up by the PMC (blue arrow highlights an example).

### Visual assessment of PMC and NAVcor performance in the younger cohort

Figure 4 shows example R1 maps from a single individual of the younger cohort showing effects of different combinations of *PMC on/off* and *NAVcor on/off*. The figure highlights regions in the original R1 maps (Figure 4, top row) that are noticeably improved by both NAVcor and/or PMC and their combination. Maps of the differences between the navigator corrected and original R1 maps (Figure 4, bottom row) show the distribution and size of NAVcor corrected effects. PMC reduced the level of motion artifacts and increased local GM/WM contrast. Depicted maps are a representative example for the cohort showing the most typical features of the artifacts. However, the extent and the location differed from one participant to another. To ensure an unbiased comparison, the selected participant had the most similar amount of head motion throughout the *PMC on* and *PMC off* measurements. The averaged integrated motion metric (59) for this volunteer from PD and T1-weighted acquisitions was 20.6 mm and 19.4 mm for *PMC on* and *PMC off* respectively, which was less than the average values across the whole group (presented as [mean ± standard deviation]: (43±22) mm and (48±31) mm respectively).

**FIGURE 4.**
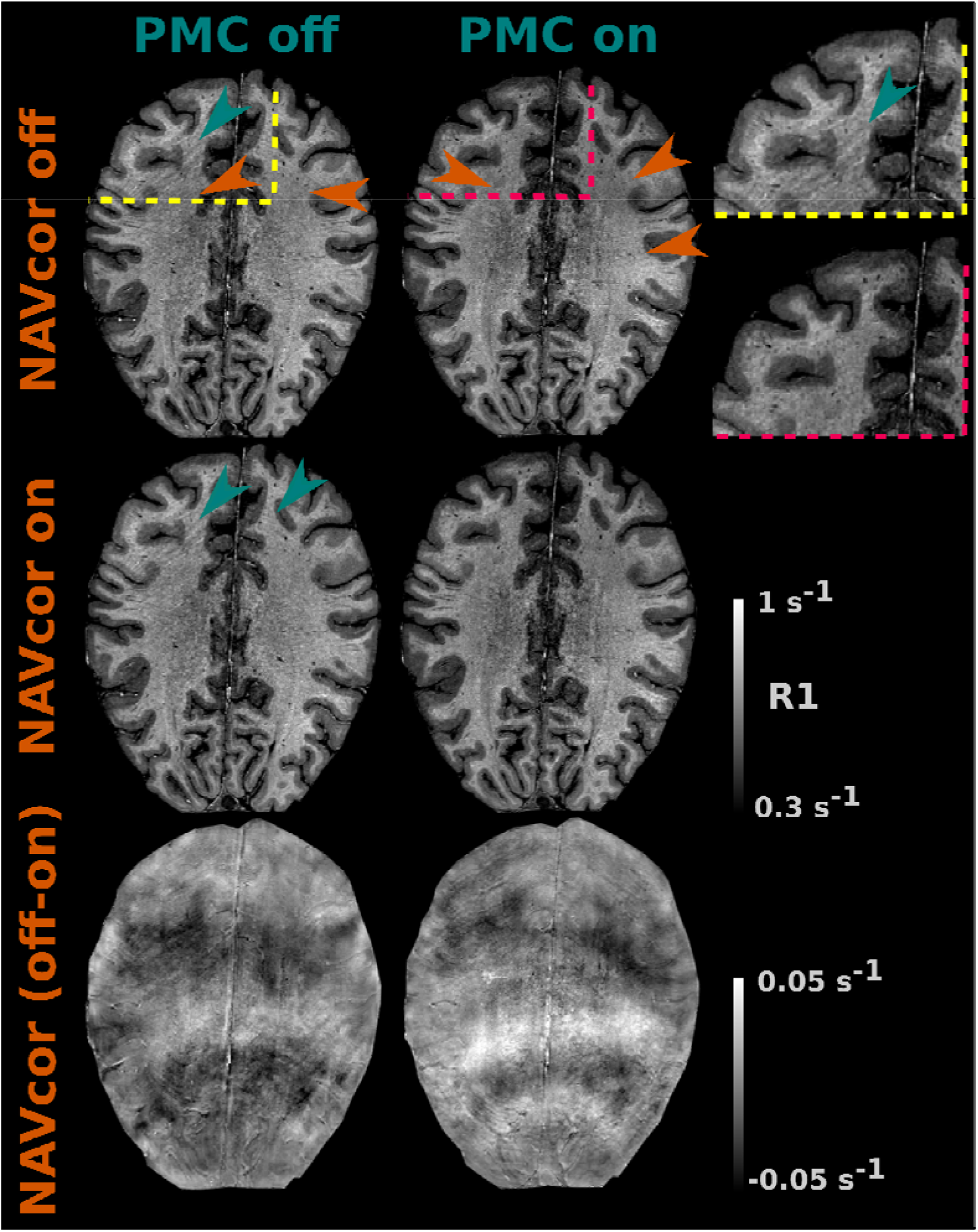
R1 maps of a compliant volunteer from the younger cohort demonstrating the impact of the individual and combined corrections for B0-fluctuation (NAVcor) and motion (PMC). Top row: original R1 maps acquired without (left column) and with (middle column) PMC. Middle row: R1 maps after application of NAVcor. Bottom row: difference maps of original minus NAVcor R1 maps highlighting the regions where NAVcor had the highest impact. Sections (zoomed into frontal areas of left and middle column) in the top right corner (top row, right column) contrast motion artifacts in PMC off (yellow border) versus PMC on (red border) conditions. The color-coded arrows point at the areas where corresponding artifacts are apparent (NAVcor in orange and PMC in turquoise).

### Quantitative assessment of spatial inhomogeneities in both cohorts

In general, the PMC and NAVcor reduced the level of spurious spatial inhomogeneities. We observed a higher variability of MR parameters in GM than in WM (orange vs green boxplots in Figure 5). In the younger cohort, the average relative reduction in inhomogeneity CoV between the NAVcor on and off conditions for all parameter maps was (mean ± SD across group): (1.9±1)% for PMC off and (2.1±1.5)% for PMC on in WM and (1.0±0.6)% for PMC off and (1.0±1.2)% for PMC on in GM. In the elderly and patient cohort, the relative reduction in inhomogeneity CoV was (9.2±2.1)% in WM and (5.9±2.3)% in GM, suggesting that the NAVcor is more relevant in this group.

**FIGURE 5.**
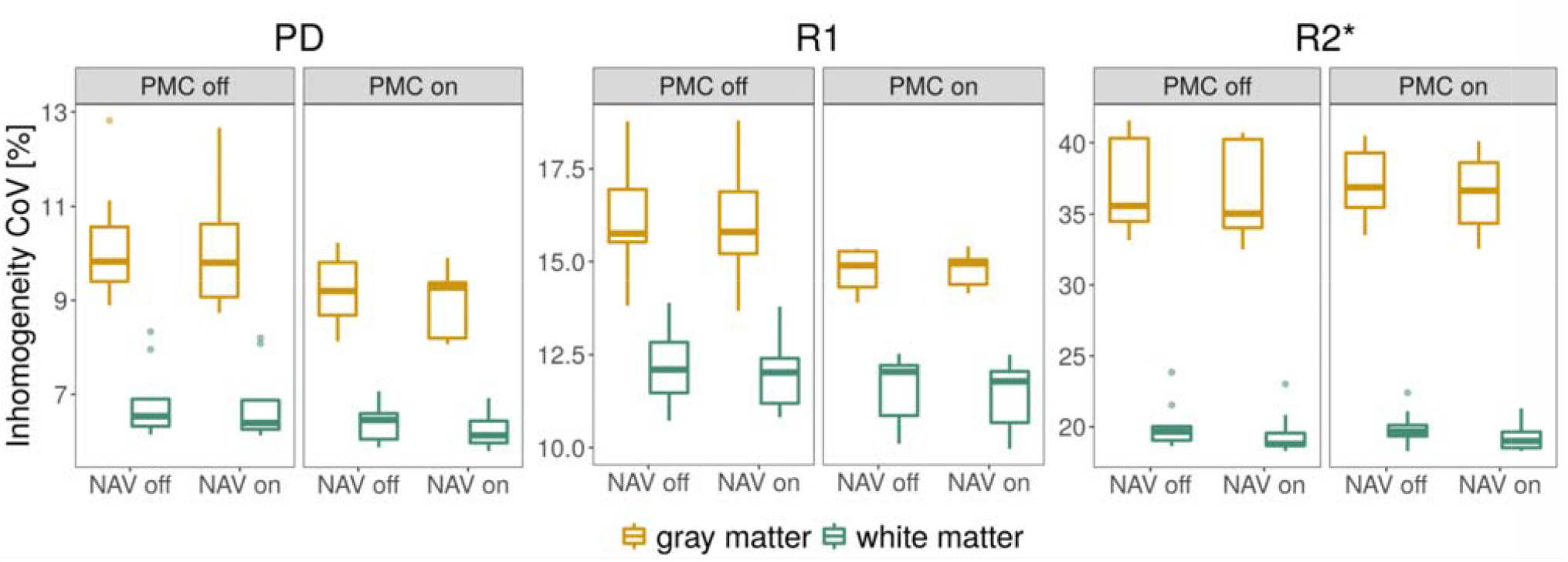
Inhomogeneity CoV within GM (yellow) and WM (green) of the younger participant group extracted from PD, R1 and R2* maps that were either corrected for rigid motion and dynamic B0-fluctuations (PMC on, NAV on) or not (PMC off, NAV off). The box plots contain 11 datasets from 10 participants (two datasets from one participant). Boxplots indicate median value, first and third quartiles and outliers.

Statistical analysis was used to determine if there is an interaction between the two corrections in the younger cohort. Two-way ANOVA of 11 datasets from 10 subjects (1/10 subjects was invited twice) was performed for all three MR parameters. The effect of PMC did not reach significance for any parameter within either WM or GM (all p > 0.5). The NAVcor effect was significant in two cases: p = 0.02 for PD in GM and p = 0.009 for R2* in WM. The interaction effect between PMC and NAVcor was not significant for any of the parameters (all p > 0.12).

### Visual assessment of parameter maps in elderly and patient cohort

Example parameter maps of dementia patients without (left column) and with (middle column) NAVcor are displayed in Figure 6 together with difference maps highlighting areas affected by dynamic B0-fluctuation-related artifacts (right column). The effects of correction were observed on PD, R1 and R2* maps. The variety of artifact types that were improved with NAVcor included regional changes of signal level visible in all maps, blurring in the left subthalamic nucleus and putamen in the R2* map and localized fluctuating signal bands that were evident in the parietal lobe in the R1 map as well as frontal region in the R2* map. In this experiment, data were always acquired with PMC turned on, i.e. in the *PMC on* condition. The average integrated motion metric within the elderly and patient group was (98±53) mm (mean and SD across all participants and acquisitions).

**FIGURE 6.**
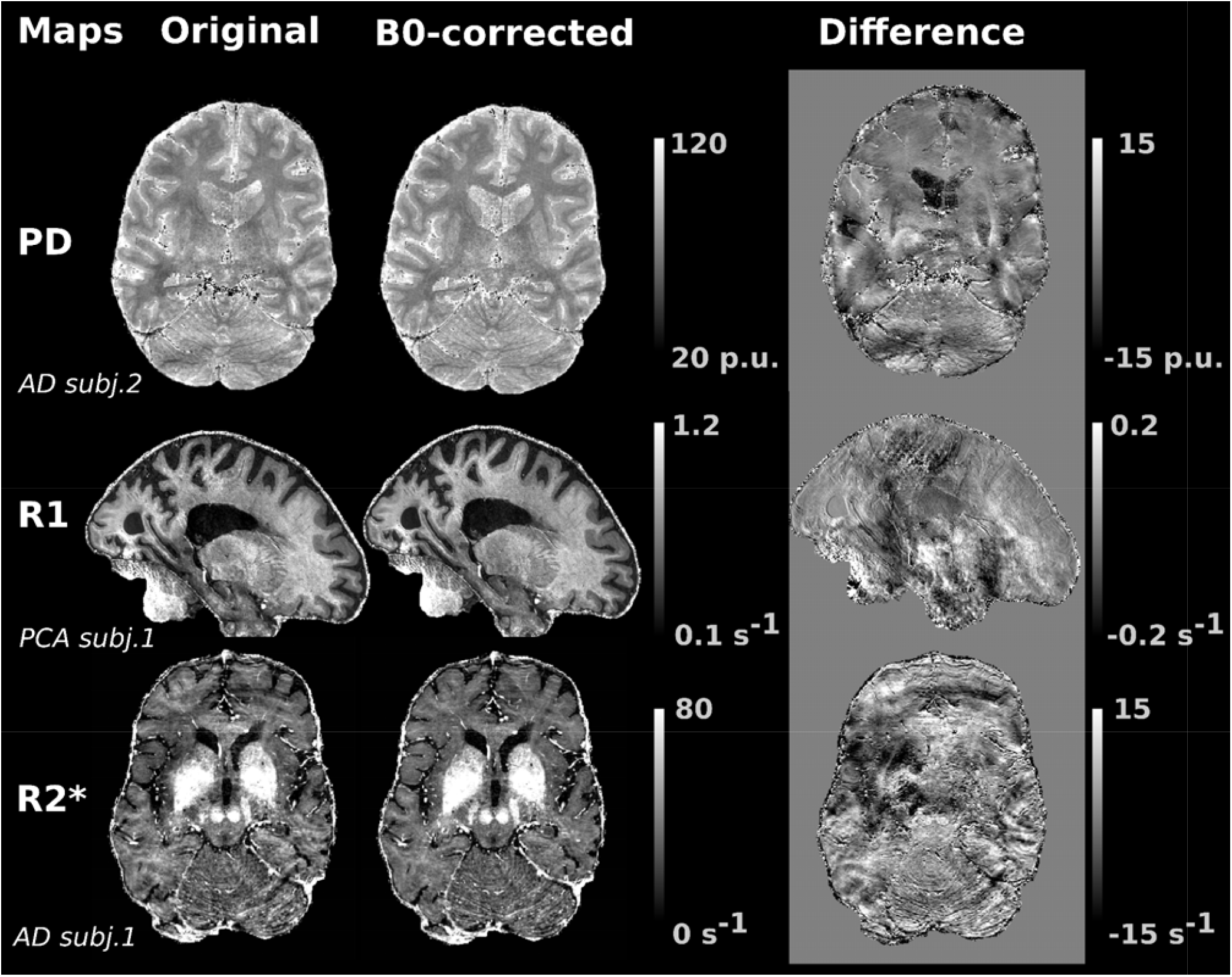
Example parameter maps of two AD patients (PD, R2*) and one PCA patient (R1) show the range of effects of NAVcor on the quality of maps. Difference maps of original minus corrected maps (right column) visualize the location and artifact types, such as regional signal fluctuations (darker and lighter areas spread out over larger brain regions in all map types), ripple artifact (parietal lobe in R1, frontal lobe in R2*) and blurring (left basal ganglia in R2* and PD).

The navigator correction was also applied to the in-line SENSE auto-calibration reference lines. Thus, we occasionally observed reductions in residual aliasing, i.e., SENSE ghost, due to reduction of motion/B0 effects in the in-line reference data (Figure 7).

**FIGURE 7.**
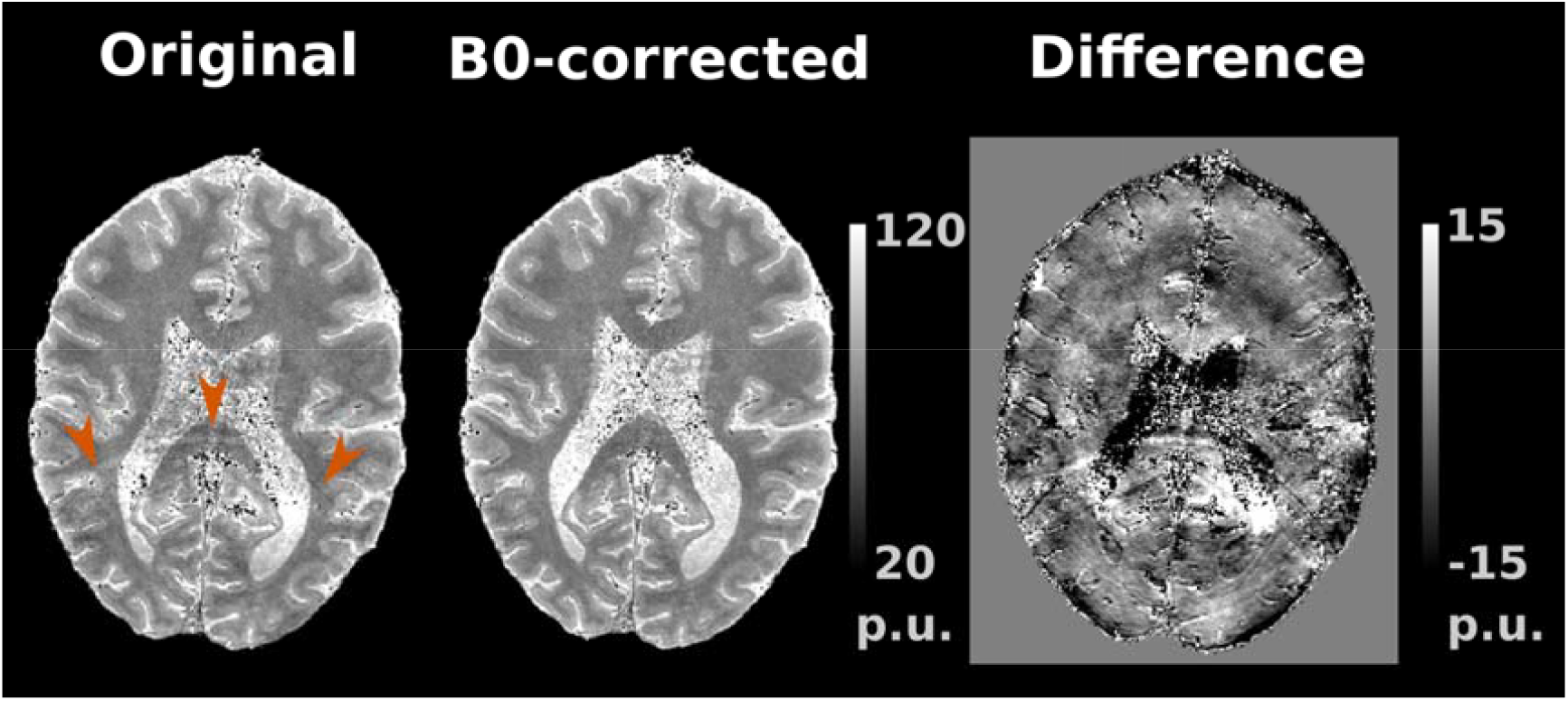
SENSE unfolding artifact reduced by NAVcor in a PD map of a PCA patient. The arrows and the difference map highlight the semicircular SENSE ghost (left and right), which is not visible in the NAVcor PD map (middle).

### Scan-rescan reproducibility of parameter maps in the elderly and patient cohort

Parameter maps from two different sessions were used to determine the scan-rescan reproducibility with and without NAVcor. Figure 8 shows the average scan-rescan CoV for the GM and WM ROIs for all participants and maps, superimposed by the mean over participants (darker colors). On average, the scan-rescan CoV is lower for *NAVcor on* compared to *NAVcor off* in line with an increased reproducibility of the parameter maps when NAVcor was applied. The improvement in scan-rescan CoV computed as a relative change and averaged over the three map types was (14.7±9.0)% in WM and (11.9±12.0)% in GM regions. The improvement in reproducibility was observed in 55/60 analysis cases (10 volunteers x 3 map types x 2 tissue classes). Only 5 cases showed a decrease in reproducibility on average by 9% (mainly driven by a single outlier: 3.6%, 3.5%, 0.7%, 5.6% and 31.1%). A one-tailed t-test was performed and showed significance for all three contrasts both in WM (all p≤0.004) and GM (all p≤0.025).

**FIGURE 8.**
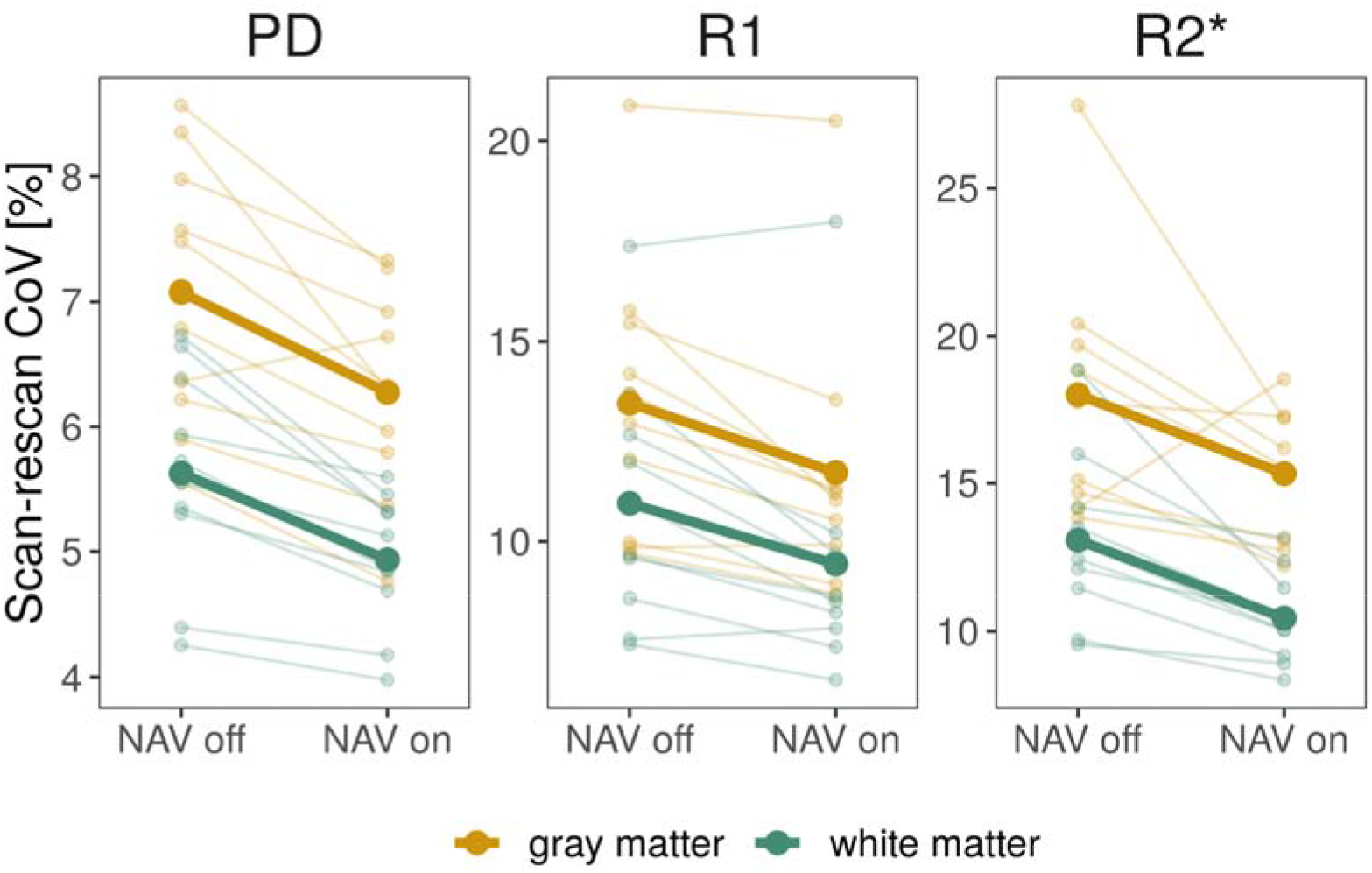
Scan-rescan reproducibility of all participants’ PD, R1 and R2* parameter maps assessed within white matter (in green) and gray matter (in yellow) separately. Average scan-rescan CoVs in GM and WM ROIs are depicted for each map type and overlaid by the mean over participants in thick darker lines of the respective color.

## DISCUSSION

We developed a method for correcting dynamic B0 field fluctuations and rigid-body head motion in multi-echo 3D GRE acquisitions at 7T based on FID navigators and optical PMC. We demonstrated an improvement in the quality and scan-rescan reproducibility of high-resolution quantitative R1, PD and R2* maps. The improvements were observed in healthy controls but most prominently in a group of patients with dementia and elderly controls with consistent reductions of scan-rescan CoV > 10%. The data in healthy young volunteers showed that the navigator correction and PMC are complementary and result in an additive correction effect, i.e., they are best used in concert.

The correction of physiological artifacts including dynamic changes of B0 and head motion is essential for sub-millimeter resolution scans at 7T, since long scan times >20 mins are required and even involuntary movements easily exceed the voxel size (cf. Figure 3). Moreover, the high static B0 field results in higher absolute dynamic B0 fluctuations. The consequent artifacts reduce the image quality and effective resolution (23,38) and hinder important studies on the mesoscopic brain structure and microstructure in health and disease (2).

The quantitative mapping of multiple physical parameters is an effective approach to non-invasive microstructure imaging, since the different parameters are differently sensitive to the underlying brain microstructure (2). The MPM method can efficiently acquire PD, R1 and R2* maps and has been widely used to study microstructure in healthy volunteers (6,13,16,60,61) and patients (e.g., (20,62)). Its potential for resolving thin intra-cortical layers and U-fibers has been demonstrated (6,12,13) but remains limited due to physiological artifacts.

Dynamic B0-fluctuation correction has been mainly exploited for acquisitions particularly vulnerable to B0 variations due to long echo times, such as T2*-weighted imaging and mapping (22,63,64), including susceptibility weighted imaging (SWI) and quantitative susceptibility mapping (QSM) (65). In this work, we extended the use of PMC and B0-corrections to R1 and PD in addition to R2* maps and showed that they improved all three types of parameter maps. Since these parameters are related to myelination (60,61,66), they are important markers complementary to the iron sensitive R2* and QSM in neuroimaging of healthy cohorts (2,13,60,61) and pathology, neurodegeneration and trauma (20,67,68).

To reduce the physiological artifacts, we implemented PMC and navigator corrections in the multi-echo 3D GRE sequence used for MPM allowing for correction of artifacts caused by motion and dynamic B0 changes. In our implementation, the navigator was acquired after the imaging echoes (65) in the absence of gradient encoding. In comparison to previous studies, which used multi-echo readouts, and collected FIDs right after the excitation pulses (46,69) the FID had a lower SNR and was affected by eddy currents due to multi-echo readout. On the other hand, this approach increased the phase contrast and could preserve minimal TEs. We increased the TR to reduce eddy current induced phase variations affecting the FID navigator signal and only used part of the acquired FID signal (Figure 1). These eddy current specific settings may need to be adjusted when the readout gradients and their timings are changed.

Typically, the SNR and quality of the FID navigators was sufficient to estimate the dynamic B0 changes. However, in a minority of cases, some RF channels exhibited navigator time series with low SNR, which resulted in unreliable phase estimates, unstable navigator correction and image artifacts. Exploration of instabilities suggested that often RF channels were affected whose receive coils covered areas with high B0 field inhomogeneities such as basal brain areas, which is in line with undesired dephasing effects across the relatively large receive volumes resulting in low signals. Other navigator correction approaches, which work on slice-based 2D GRE acquisitions (21,40) instead of 3D GRE or use spatially encoded navigators (65), are less sensitive to the dephasing effects, since the excited or encoded navigator voxel volumes are smaller. Furthermore, we observed that motion visible on PMC traces often coincided with the onset of low navigator SNR periods. We hypothesize that motion of tissue relative to the rest of the head (e.g. jaw) could be an underlying cause, but the issue was also noticed in channels that were not located close to the face or shoulders.

To improve the robustness of the phase estimation from the FID navigator signals, we developed a temporal low pass filter and regularization, which adapted to the local SNR, and an additional consistency check across RF channels. The adaptive smoothing exploits the temporal smoothness of physiological processes, enhancing the SNR while minimizing the loss in temporal resolution in the navigator time series. It is complemented by a check for consistency across RF channels and correction by replacing with a modeled signal where necessary, exploiting the spatial smoothness of the dynamic frequency shifts. Both processing steps and their free parameters are automatically adjusted to each dataset, significantly improving the robustness of the navigator correction.

The navigator-based dynamic frequency measurements reliably detected respiratory dynamics and different motion effects in line with breathing sensor and PMC measurements (Figure 3). Average peak-to-peak field shifts during the respiratory cycle in young adults were (1.6±0.5) Hz, which is in agreement with previous studies at 7T (9,10,45). The frequency shifts in elderly controls and patients with dementia were higher (on average 2.7 Hz) but lower than the average 6 Hz reported previously (21). Global frequency drift during the measurement, as a result of both hardware and physiological changes, was on average 24 (4 times higher than in the younger cohort) over the course of the ∼20 mins long measurements.

The effectiveness of PMC and navigator correction was corroborated by qualitative and quantitative analysis. Visual inspection of the parameter maps revealed a rich set of artifacts ranging from some localized motion artifacts to smooth signal variations reduced by the navigator correction in line with previous reports ((35); Figure 4).

The correction performance was quantified by measuring the spatial homogeneity of parameter maps and also their scan-rescan reproducibility within WM and GM. WM and GM masks were derived from SPM12’s unified segmentation tissue probability maps and thresholded conservatively to reduce partial volume effects and edge artifacts. The improvement in homogeneity CoV due to the navigator correction was on average approximately 5-fold higher in the elderly volunteers and dementia patient cohort than in the younger volunteers group. This is likely due to the higher level of artifacts in this population, which is driven by increased head motion, different types of motion and breathing patterns (21,27). We note that the spatial inhomogeneity metric not only captures artifactual but also biological variation even within a single tissue class. Thus, the metric is best regarded as an upper estimate of variation and is expected to underestimate the effectiveness of the corrections.

The scan-rescan reproducibility metric is much less affected by biological variations, since baseline differences are disregarded when looking at differences between two scans, but it requires two scans instead of a single scan. Another advantage is that it simulates longitudinal imaging studies, which are frequently performed in developmental, aging or intervention studies. Thus, the results can assist power analyses for these studies (4). For example, the 13% reduction in scan-rescan CoV (Figure 8) due to the navigator correction in the group of dementia patients and elderly volunteers is comparable to the sensitivity increase that would be expected for extending the scan time by ca. 27% (under the simplifying assumption of stationary Gaussian noise). This is particularly important when targeting small effects such as ∼1% annual changes in R2* in the basal ganglia in Alzheimer’s disease patients or cortical R1 in Parkinson’s disease patients (14,70).

Even in the challenging dementia patient and elderly subject group we observed an increase in scan-rescan CoV in only 8% of the cases (5 cases out of 60). This is similar to a previous study which reported a decrease in NAVcor image quality in 1 out of 10 AD patients (21). These cases may be identified by visual inspection or automated checks and the image reconstruction repeated without navigator correction.

PMC and B0-correction are expected to be complementary and their improvements to be additive. PMC updates the geometry of the scan based on a rigid body assumption, without taking into account movements such as brain tissue pulsations. It also does not correct for phase errors caused by motion of susceptibility interfaces (42), which is where navigator correction comes in. However, floor/ceiling effects in their performance or interactions between susceptibility-related dynamic changes and motion may result in an interaction between the two corrections. Statistical testing on an inhomogeneity measure did not reveal any significant interaction between the two corrections, supporting that both are additive and may be applied simultaneously. However, the testing was performed in the younger group for which there was no significant influence of PMC, since the non-factorial experimental design in the elderly cohort was not suitable to study these effects.

Various methods for motion and B0 corrections have been developed. Dynamic shimming approaches vary the shim currents to compensate for time varying B0 changes (10,35,71), which directly compensates higher order spatial B0 distortions. Concurrent field monitoring can be used to inform the dynamic shimming (35) or post-processing corrections (44). However, compared to navigator-based corrections, these methods require additional hardware and integration into the MR system.

Alternative types of motion correction methods include fat navigators (33,34), self-navigated methods (31), navigator echoes (32), other external tracking devices (35) or modeling motion in the image reconstruction (72). We opted for the use of optical tracking and PMC, since it does not require significant changes in the pulse sequence nor the scan duration unlike additional navigator scans and it corrects the data during acquisition. The majority of approaches including PMC correct for rigid body motion only and ignore interactions of motion with the receive field. Particularly when using multi-channel head coils with small RF coils, motion of the head through the receive field and the consequent signal fluctuations become more relevant. At very high resolutions also brain pulsations and elastic movement become more relevant.

While the navigator correction in combination with PMC is important to achieve low artifact levels in elderly volunteers and patient groups, the lower homogeneity CoV in the younger volunteers group suggests it may not be as critical in younger healthy cohorts. Care should be taken when extrapolating the study results to other cohorts and imaging protocols, since the duration of scans, types of motion and hardware setup may significantly impact artifact levels and correction performance.

## CONCLUSIONS

We developed a method for correcting dynamic field fluctuations and rigid-body head motion at 7T based on navigators and optical PMC, which led to a significant improvement in the quality and scan-rescan reproducibility of quantitative high-resolution R1, PD and R2* maps. The improvements were most striking in dementia patients and elderly healthy controls. Thus, we recommend including the corrections in studies of such cohorts and for 3D imaging with long acquisition times, high spatial resolutions and when targeting small effects.

## Data Availability

All data produced in the present study are available upon reasonable request to the authors.

https://github.com/lenkavaculciakova/NavCorPMC

## ACKNOWLEDGEMENTS

We would like to thank P. Scheibe and D. Hänelt (Department of Neurophysics, Max Planck Institute for Human Cognitive and Brain Sciences) for their help in early stages of the study with visual inspection and rating of the acquired data. We would like to thank C. Rüger (Department of Cariology, Endodontology and Periodontology, University Medical Center Leipzig) for manufacturing the mouth guards for optical prospective motion correction. We also thank K. Lancaster, D. Davidson and N. Hunt (UCL Eastman Dental Institute, London, UK) for manufacturing the mouth guards in the elderly and dementia patient group. The research leading to these results has received funding from the European Research Council under the European Union’s Seventh Framework Programme (FP7/2007-2013) / ERC grant agreement n° 616905, and from the NISCI project funded by the European Union’s Horizon 2020 research and innovation programme under the grant agreement No 681094, and the Swiss State Secretariat for Education, Research and Innovation (SERI) under contract number 15.0137. NW received funding from the BMBF (01EW1711A & B) in the framework of ERA-NET NEURON. TV is funded by an Alzheimer’s Research UK (ARUK) PhD scholarship (ARUK-PhD2018-009). The Wellcome Centre for Human Neuroimaging is supported by core funding from the Wellcome [203147/Z/16/Z].

